# Integrating environmental sustainability in clinical counselling: a randomised, double-blinded, experimental vignette study of satisfaction with care

**DOI:** 10.1101/2025.03.23.25324468

**Authors:** Egid M van Bree, Laurens C van Gestel, Eva H Visser, Jiska J Aardoom, Evelyn A Brakema, Marieke A Adriaanse

## Abstract

**STRUCTURED ABSTRACT:** *Objective:* to explore whether advising environmentally sustainable treatment options in clinical counselling affects patients’ satisfaction with care compared to less sustainable (standard) options – accounting for differences in severity and types of medical problems.

*Design:* a randomised, double-blinded, experimental vignette study with a four (between-group, Type of Advice) x two (between-group, Severity) x five (within-group, Medical Problem) mixed design. Participants received short descriptions of hypothetical patient-physician interactions based on their group allocation and subsequently indicated their satisfaction with care.

*Setting:* a general practice or hospital visit.

*Participants:* a representative sample of the general Dutch adult population.

*Interventions:* four different types of medical advice, varying in level of environmental sustainability and whether sustainability is mentioned explicitly.

*Main outcome measures:* satisfaction with care, operationalised as: treatment acceptability, trust in the physician, trust in the treatment, and the feeling that the physician prioritised their health (Likert scales, 1=strongly disagree, 7=strongly agree).

*Results:* 1,536 participants completed the study. Across vignettes, participants receiving the less sustainable advice (M=5.6, SD=1.2) were generally more satisfied than participants receiving the more sustainable types of advice (P’s<.001). Participants receiving the explicitly more sustainable advice (M=4.8, SD 1.6) were generally less satisfied than participants receiving the implicitly more sustainable types of advice (P’s<.001). Differences were larger for the high-severity conditions (mean differences 0.4 to 1.8, P’s<.001), non-significant for most low-severity conditions, and varied per medical problem.

*Conclusions:* advising more sustainable treatment options for high-severity conditions may negatively affect patients’ satisfaction with care, especially when sustainability is mentioned explicitly. However, the presence and size of the observed effect varied across medical problems and was generally non-significant or small for low-severity conditions. Clinicians’ situational awareness may differentiate whether sustainability should be discussed, or only guaranteed through institutional level decisions.

## INTRODUCTION

The healthcare sector faces an urgent challenge to reduce its environmental footprint.(1) Especially in high income countries, healthcare systems contribute notably to national CO_2_-emissions (6-8%) and natural resource use (13%).(2,3) To date, over 90 countries pledged to deliver their care more environmentally sustainable.(4) Concurrently, a rapidly growing body of literature supports the inception of evidence-based, sustainable decision-making.(5)

Including environmental sustainability as a quality criterion for care delivery is frequently suggested as a key strategy for its incorporation in healthcare.(6,7) Consequently, clinicians are called upon to consider the environmental impact of healthcare when choosing between treatment alternatives and designing clinical guidelines.(8,9) In the case of clinical equipoise, the more sustainable treatment option is preferred. However, expecting patients to respond negatively when (explicitly) considering the environmental option in clinical counselling may be perceived as a barrier to do so.(10,11)

A recent experimental vignette study concluded that primary care patients’ satisfaction with care was not affected when an equally effective, sustainable option was advised for low-severity health complaints.(12) When environmental sustainability was also explicitly mentioned as an argument for advising the sustainable option, effects (positive/negative) varied per health complaint. This suggests that worries regarding negative patient responses to more sustainable treatment options may not be justified. To date, patients’ satisfaction with sustainable treatment options for more severe health complaints remains largely unknown.

That is, of the studies suggesting that patients may be willing to consider the environment in their treatment choices,(13-16) three studies also indicated that willingness is lower for more severe medical conditions.(13,16,17) However, all of these studies directly inquired patients’ willingness and may not necessarily reflect their responses when exposed to (hypothetical) clinical counselling. Moreover, evidence regarding the effect of explicitly counselling patients on the environmental impact of treatment options is limited to the aforementioned primary care study.(12)

Considering these gaps in the literature, the present study primarily aims to explore whether advising environmentally sustainable treatment options with or without explicitly mentioning sustainability as an argument in clinical counselling affects patients’ satisfaction with care, compared to less sustainable (standard) treatment options. Furthermore, it investigates whether a possible effect is influenced by the perceived severity of the medical problem and/or the type of medical problem.

## METHODS

### Study design

We performed a randomised, double-blinded experimental vignette study. We used an online survey tool to provide participants with short descriptions of hypothetical patient-physician interactions.

Next, participants answered a corresponding set of questions which served as our dependent variable. The mixed experiment had a 4 (between-group, Type of Advice) x 2 (between-group, Severity) x 5 (within-group, Medical Problem) design with satisfaction with care as dependent variable (Figure 1). The study design is a conceptual replication and extension of the aforementioned primary care study.(12) We investigated a different set of medical problems in a general practice setting and added more severe medical problems in a hospital setting for variance in severity. We preregistered the study and its planned analysis on AsPredicted (aspredicted.org/mhzn-pmv6.pdf) and collected data between the 16^th^ and 31^st^ of May 2024. The authorised departmental review committee of the Leiden University Medical Center gave ethical approval for the conduct of the study (#24-3033). Participants gave informed consent prior to participation. All study materials are available via the Open Science Framework (<link>).

**Figure 1.**
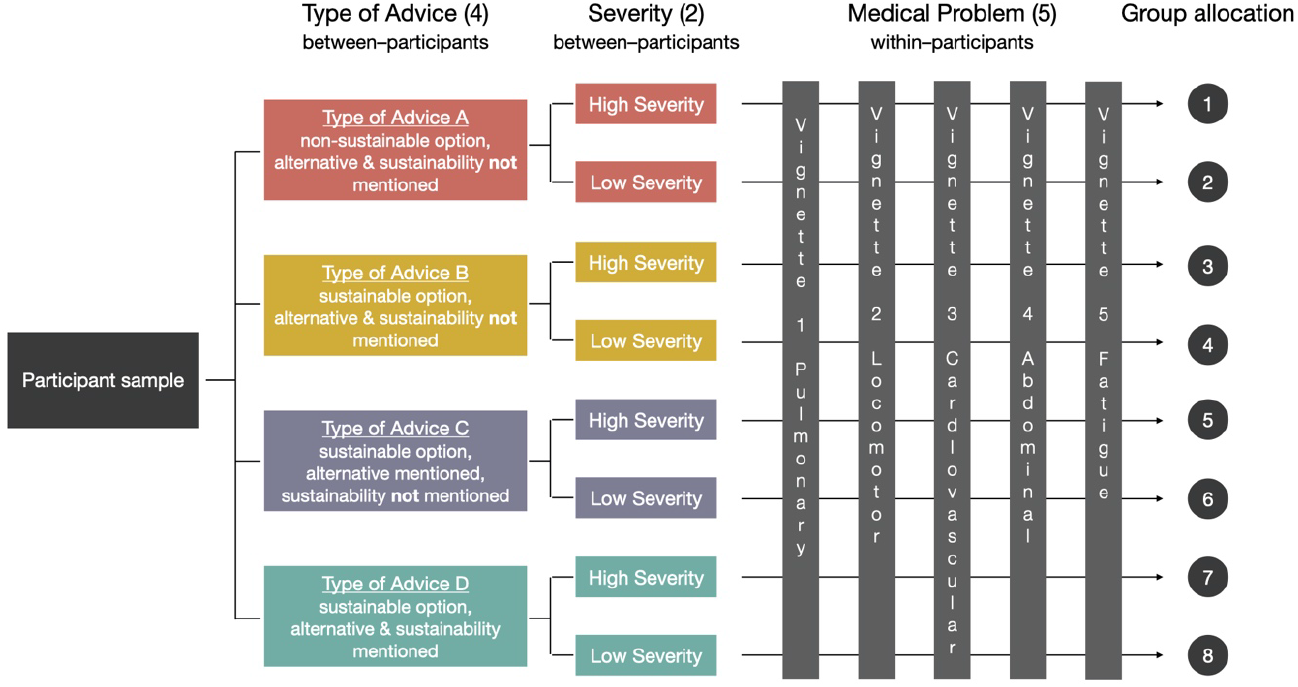
Flow diagram of the study design and participants’ group allocation

### Participants and procedure

We calculated a required sample size of 1,456 participants (*n*=182 per group) based on an a priori power calculation (α=.05, power=.8) for a smallest effect size of interest of mentioning environmental sustainability on participants’ satisfaction with care using G*Power 3.1. We recruited a representative sample (based on sex, age, education level, and geographical distribution) of the general Dutch adult population, able to read and understand the Dutch language. To facilitate access to and quality of our sampling population, we collaborated with a research agency (Flycatcher.eu) whose online panel is ISO-20252 certified for quality requirements to conduct scientific research.

Participants were invited to participate via email, with a maximum of two reminders if participation had not been initiated or completed, and received a compensation from the research agency upon completion (equivalent to €2.60).

We automatically blinded, randomised, and allocated participants to one of the eight study groups in the online software. The order in which the different vignettes were shown varied across participants. Enrolment continued until the minimum required number of participants per group had been reached. To avoid selection bias, we concealed the exact aim of the study before participation and only informed participants that the research was intended to inquire their satisfaction with hypothetical doctors’ visits. Participants could end their participation at any moment and a debriefing of the actual research aim and opt-out possibility were included at the end of the study.

Participants who did not meet the survey’s attention check or had poor response quality (as preregistered) were excluded from the analyses.

### Materials

We carefully constructed text-based vignettes in B1-level Dutch, in line with published guidance on vignette validity.(18) All vignettes followed the same structure to describe a hypothetical patient-physician interaction for each of the five Medical Problems based on the participant’s group allocation. We did not use supporting pictures or videos, considering our focus on the content of the counselling. Low-severity scenarios described a general practice setting and high-severity scenarios a hospital setting where the patient had been referred (Boxes 1-2). Note that in the Netherlands, a referral from primary care is required for consultations in the hospital. Medical problems related to different origins, including pulmonary (asthma/lung cancer), locomotor (sprained/broken ankle), cardiovascular (hypertension/angina), abdominal (cholecystolithiasis/appendicitis), and fatigue problems (possible hypothyroidism/malignancy).

Scenarios were directly followed by a corresponding Type of Advice, drawn up in close consultation with relevant medical specialists and general practitioners. Available options were: A) the standard, environmentally less sustainable treatment (“Less Sustainable”); B) the environmentally more sustainable treatment (“Sustainable”); C) the environmentally more sustainable treatment, while mentioning that there is an alternative (“Sustainable with Alternative”); and D) the environmentally more sustainable treatment, while mentioning that there is an alternative and that the suggested option is more environmentally sustainable (“Sustainable made Explicit”). From a medical perspective, the quality of care and patient’s health outcome were considered equal for the Less Sustainable and Sustainable options – nuances left aside to benefit readability.

We pilot-tested the vignettes’ face validity. First, we consulted two physicians not involved in the research team and two laypersons of different education levels to freely comment on the readability and credibility of vignettes – and adjusted accordingly. Second, we asked members of the general Dutch population of a different online panel (*n*=85) to rate severity on two seven-point Likert scales (severity and urgency) and comment on readability. For all vignettes included in the final study, the high-severity version was indeed rated as more severe than the low-severity version. A description of all vignettes is provided in Supplement A and details regarding the pilot study in Supplement B.

#### Box 1. Translated example of a low-severity vignette including different types of advice

**Vignette 1: pulmonary (asthma)**

<u>Scenario:</u>

You are visiting your general practitioner because you have a cough and feel somewhat short of breath for a couple of weeks. After the conversation and an examination, the general practitioner concludes that you have asthma. The general practitioner discusses with you what to do next.

<u>Less Sustainable advice:</u>

He/she proposes to prescribe you a metered dose inhaler, to make sure that you don’t become short of breath again.

<u>Sustainable advice:</u>

He/she proposes to prescribe you a dry powder inhaler, to make sure that you don’t become short of breath again.

<u>Sustainable with Alternative advice:</u>

Different types of medication are available: 1) a metered dose inhaler, or 2) a dry powder inhaler. He/she proposes to prescribe you the dry powder inhaler, to make sure that you don’t become short of breath again.

<u>Sustainable made Explicit advice:</u>

Different types of medication are available: 1) a metered dose inhaler, or 2) a dry powder inhaler. He/she proposes to prescribe you the dry powder inhaler, to make sure that you don’t become short of breath again and because this medication is less burdensome for the environment than the metered dose inhaler.

#### Box 2. Translated example of a high-severity vignette including different types of advice

**Vignette 1: pulmonary (lung cancer)**

<u>Scenario:</u>

Your general practitioner has referred you to the hospital because you have been coughing a lot and feeling tired for a couple of weeks. After the conversation and an examination, the physician concludes that you have early-stage lung cancer. The physician discusses with you what to do next.

<u>Less Sustainable advice:</u>

He/she recommends surgery to remove the sick part of your lung.

<u>Sustainable advice:</u>

He/she recommends targeted radiation to kill the cancer cells.

<u>Sustainable with Alternative advice:</u>

Two treatments are possible: 1) surgery to remove the sick part of your lung, or 2) targeted radiation therapy to treat the sick part of your lung.

He/she recommends targeted radiation to kill the cancer cells.

<u>Sustainable made Explicit advice:</u>

Two treatments are possible: 1) surgery to remove the sick part of your lung, or 2) targeted radiation therapy to treat the sick part of your lung.

He/she recommends targeted radiation to kill the cancer cells, also because radiotherapy is less burdensome for the environment than a surgery.

### Outcome measures

#### Satisfaction with care

We operationalised satisfaction with care as the mean of four statements, inquired directly after participants’ exposure to a vignette: “I agree with this treatment”, “I trust this physician”, “I trust this treatment”, and “this physician pays attention to my health”. Participants rated items on seven-point Likert scales (1=strongly disagree, to 7=strongly agree). In addition, they received an open-ended question after the four items, in which participants could indicate the most important question they might have for the physician after receiving their advice (reported in Supplement C).

#### Sample characteristics

Prior to presenting the vignettes, we inquired about participants’ type of living area, self-rated health status, and baseline trust in physicians. Health status was operationalised as a single, five-point item of the SF-36 Health Survey’s Dutch version.(19) We measured baseline trust in physicians on a five-point Likert scale. Demographics were collected automatically from the online panel database (sex, age, and education level). After completing the vignettes (to avoid influencing our dependent variable), we asked participants to indicate any personal experience with the Medical Problems and whether they believed in climate change. For exploratory purposes, all participants responded to three statements inquiring their view on sustainable healthcare and three statements related to climate change – if they indicated to believe in it (reported in Supplement D). Participants rated statements using five-point Likert scales (1=strongly disagree, to 5=strongly agree).

### Analysis

We analysed data using R v4.4.1. We calculated and reported a descriptive sample overview using frequencies, means (M), and standard deviations (SD) for the dependent variable and demographics. The dependent variable consisted of a composite score for the satisfaction with care statements (Cronbach’s *α*=.93 to .96, Supplement E). Furthermore, we performed a randomisation check for baseline characteristics, personal experience with the medical problems, and belief in climate change using Chi-Square and Kruskal-Wallis tests.

To answer the main research questions, we performed the preregistered three-way Type of Advice (between) x Severity (between) x Medical Problem (within) mixed ANOVA on satisfaction with care. Main effects were presented as estimated marginal means, Huynh-Feldt corrected F-test values, and effect sizes (*η*_*p*_^*2*^). We decomposed main effects and two- or three-way interactions via exploratory post-hoc comparisons using Bonferroni corrections, reporting results as mean differences per Medical Problem and Type of Advice including 95% confidence intervals (CI). As preregistered, we performed aforementioned analyses with and without the exclusion of outliers (defined as 3 SDs above or below the mean). As this generally did not affect the results, we only reported the findings based on the full sample.

### Patient and public involvement

No patients or public were involved in the design, conduct, reporting, or dissemination of our research.

## RESULTS

### Sample characteristics

N=2,694 participants were invited to participate, of whom 1,536 were included in the final sample size of the study. Incomplete entries (n=90) and participants who did not meet the attention check (n=28) were excluded. Baseline characteristics (Table 1) were not significantly different between participant groups (P’s>.22). Neither were participants’ previous experience with the medical problems in the vignettes or their belief in climate change (P’s>.10), with 71.4% of participant having experienced at least one of the medical complaints (Supplement E).

**Table 1.** Participants’ baseline characteristics, stratified by group allocation.

|  | Severity, low |  |  |  | Severity, high |  |  |  |
| --- | --- | --- | --- | --- | --- | --- | --- | --- |
|  | Type of Advice |  |  |  | Type of Advice |  |  |  |
|  | LS | S | SwA | SmE | LS | S | SwA | SmE |
|  | N (%) | N (%) | N (%) | N (%) | N (%) | N (%) | N (%) | N (%) |
|  | 202 | 183 | 186 | 201 | 182 | 188 | 209 | 185 |
| Sex, female | 94 (46.5) | 97 (53.0) | 91 (48.9) | 105 (52.2) | 96 (52.7) | 92 (48.9) | 102 (48.8) | 85 (45.9) |
| Age, mean [SD] | 53.0 [17.2] | 51.1 [17.2] | 54.0 [17.0] | 49.5 [17.0] | 51.3 [17.5] | 52.3 [17.4] | 51.7 [17.4] | 51.0 [16.3] |
| Education level |  |  |  |  |  |  |  |  |
| low | 49 (24.3) | 44 (24.0) | 55 (29.6) | 49 (24.4) | 49 (26.9) | 49 (26.1) | 53 (25.4) | 48 (25.9) |
| middle | 86 (42.6) | 82 (44.8) | 75 (40.3) | 84 (41.8) | 81 (44.5) | 76 (40.4) | 103 (49.3) | 75 (40.5) |
| high | 67 (33.2) | 57 (31.1) | 56 (30.1) | 68 (33.8) | 52 (28.6) | 63 (33.5) | 53 (25.4) | 62 (33.5) |
| Living area, rural | 86 (42.6) | 77 (42.1) | 87 (46.8) | 79 (39.3) | 80 (44.0) | 72 (38.3) | 93 (44.5) | 71 (38.4) |
| Self-rated health status |  |  |  |  |  |  |  |  |
| poor | 3 (1.5) | 3 (1.6) | 9 (4.8) | 5 (2.5) | 6 (3.3) | 6 (3.2) | 5 (2.4) | 7 (3.8) |
| fair | 48 (23.8) | 40 (21.9) | 39 (21.0) | 42 (20.9) | 43 (23.6) | 38 (20.2) | 43 (20.6) | 45 (24.3) |
| good | 109 (54.0) | 99 (54.1) | 92 (49.5) | 107 (53.2) | 98 (53.8) | 102 (54.3) | 98 (46.9) | 82 (44.3) |
| very good | 38 (18.8) | 35 (19.1) | 41 (22.0) | 38 (18.9) | 31 (17.0) | 40 (21.3) | 46 (22.0) | 43 (23.2) |
| excellent | 4 (2.0) | 6 (3.3) | 5 (2.7) | 9 (4.5) | 4 (2.2) | 2 (1.1) | 17 (8.1) | 8 (4.3) |
| Baseline trust in health professional, mean [SD] | 4.1 [0.77] | 4.1 [0.83] | 4.0 [0.79] | 4.0 [0.77] | 3.9 [0.78] | 4.1 [0.71] | 4.0 [0.78] | 4.0 [0.84] |

Legend: LS = Less Sustainable, S = Sustainable, SwA = Sustainable with Alternative, SmE = Sustainable made Explicit. Baseline trust in health professional was measured on an ascending five-point Likert scale.

### Main analysis: Effect of Type of Advice, Severity, and Medical Problem on satisfaction with care

Across vignettes, satisfaction with care scores were relatively high, with all means between 4.1 [SD=1.8] and 6.1 [SD=0.9] (Table 2). The Type of Advice x Severity x Medical Problem mixed ANOVA yielded statistically significant main effects on satisfaction with care for all three independent variables (P’s<.001). For Type of Advice (F(3, 1528)=47.3, *η*_*p*_^*2*^=.09), participants receiving the Less Sustainable advice (M=5.6, SD=1.2) were generally more satisfied than participants receiving one of three Sustainable types of advice (P’s<.001). Moreover, participants receiving the Sustainable made Explicit advice (M=4.8, SD=1.6) were generally less satisfied than participants receiving one of three other types of advice (P’s<.001). The Sustainable (M=5.2, SD=1.5) and Sustainable with Alternative (M=5.1, SD=1.4) types of advice were not significantly different (P=1.00). For Severity (F(1, 1528)=30.0, *η*_*p*_^*2*^=.02), participants in the low-severity conditions (M=5.3, SD=1.4) were more satisfied than participants in the high-severity conditions (M=5.0, SD=1.5). The main effect of Medical Problem (F(3.9, 5893.3)=21.8, *η*_*p*_^*2*^=.05) indicated that, overall, the different medical problems yielded slightly different scores for satisfaction with care, with means ranging from 4.8 [SD=1.6] for the abdominal problems to 5.4 [SD=1.3] for the cardiovascular problems.

**Table 2.**
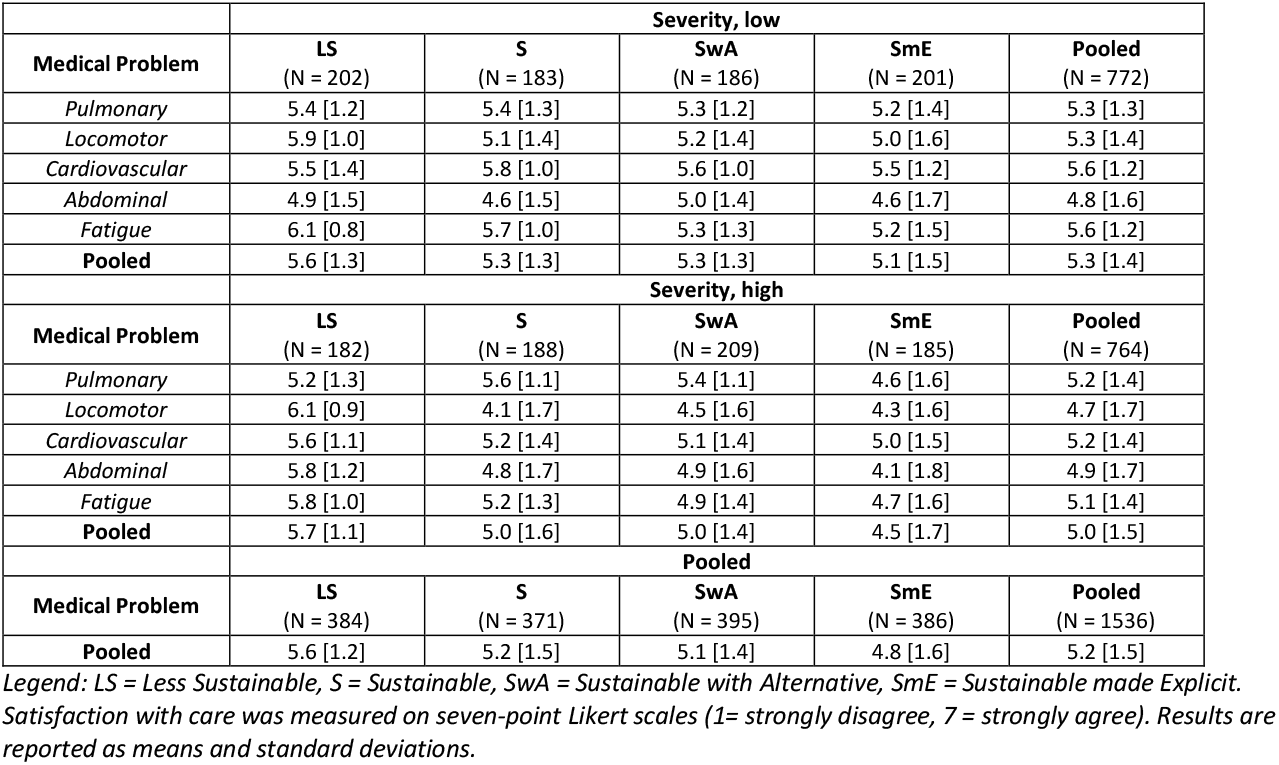
Satisfaction with care according to Type of Advice, Severity, and Medical Problem.

| Severity, low |  |  |  |  |  |
| --- | --- | --- | --- | --- | --- |
| Medical Problem | LS<br>(N = 202) | S<br>(N = 183) | SwA<br>(N = 186) | SmE<br>(N = 201) | Pooled<br>(N = 772) |
| *Pulmonary* | 5.4 [1.2] | 5.4 [1.3] | 5.3 [1.2] | 5.2 [1.4] | 5.3 [1.3] |
| *Locomotor* | 5.9 [1.0] | 5.1 [1.4] | 5.2 [1.4] | 5.0 [1.6] | 5.3 [1.4] |
| *Cardiovascular* | 5.5 [1.4] | 5.8 [1.0] | 5.6 [1.0] | 5.5 [1.2] | 5.6 [1.2] |
| *Abdominal* | 4.9 [1.5] | 4.6 [1.5] | 5.0 [1.4] | 4.6 [1.7] | 4.8 [1.6] |
| *Fatigue* | 6.1 [0.8] | 5.7 [1.0] | 5.3 [1.3] | 5.2 [1.5] | 5.6 [1.2] |
| **Pooled** | 5.6 [1.3] | 5.3 [1.3] | 5.3 [1.3] | 5.1 [1.5] | 5.3 [1.4] |
| Severity, high |  |  |  |  |  |
| Medical Problem | LS<br>(N = 182) | S<br>(N = 188) | SwA<br>(N = 209) | SmE<br>(N = 185) | Pooled<br>(N = 764) |
| *Pulmonary* | 5.2 [1.3] | 5.6 [1.1] | 5.4 [1.1] | 4.6 [1.6] | 5.2 [1.4] |
| *Locomotor* | 6.1 [0.9] | 4.1 [1.7] | 4.5 [1.6] | 4.3 [1.6] | 4.7 [1.7] |
| *Cardiovascular* | 5.6 [1.1] | 5.2 [1.4] | 5.1 [1.4] | 5.0 [1.5] | 5.2 [1.4] |
| *Abdominal* | 5.8 [1.2] | 4.8 [1.7] | 4.9 [1.6] | 4.1 [1.8] | 4.9 [1.7] |
| *Fatigue* | 5.8 [1.0] | 5.2 [1.3] | 4.9 [1.4] | 4.7 [1.6] | 5.1 [1.4] |
| **Pooled** | 5.7 [1.1] | 5.0 [1.6] | 5.0 [1.4] | 4.5 [1.7] | 5.0 [1.5] |
| Pooled |  |  |  |  |  |
| Medical Problem | LS<br>(N = 384) | S<br>(N = 371) | SwA<br>(N = 395) | SmE<br>(N = 386) | Pooled<br>(N = 1536) |
| **Pooled** | 5.6 [1.2] | 5.2 [1.5] | 5.1 [1.4] | 4.8 [1.6] | 5.2 [1.5] |

Legend: LS = Less Sustainable, S = Sustainable, SwA = Sustainable with Alternative, SmE = Sustainable made Explicit. Satisfaction with care was measured on seven-point Likert scales (1= strongly disagree, 7 = strongly agree). Results are reported as means and standard deviations.

All two-way interactions were significant (P’s<.001): Type of Advice x Severity (F(3, 1528)=47.3, *η*_*p*_^*2*^=.02), Type of Advice x Medical Problem (F(11.6, 5893.3)=26.3, *η*_*p*_^*2*^=.05), and Severity x Medical Problem (F(3.9, 5893.3)=21.8, *η*_*p*_^*2*^=.01). Given the main aims of this study, we zoomed in and decomposed only the Type of Advice x Severity interaction effect (Figure 2), by comparing the effect of the type of advice within the high- and low-severity conditions separately. Within the low-severity conditions, satisfaction with care for the Less Sustainable advice was significantly higher compared to both the Sustainable with Alternative (P=.03) and the Sustainable made Explicit (P<.001) types of advice, but not compared to the Sustainable advice (P=.11). None of the other comparisons were significant (P’s>.13). Within the high-severity conditions, however, satisfaction with care was significantly higher for the Less Sustainable advice compared to all three Sustainable types of advice (P’s<.001). Moreover, it was significantly lower for the Sustainable made Explicit advice than for the two other Sustainable types of advice (P’s<.001).

**Figure 2.**
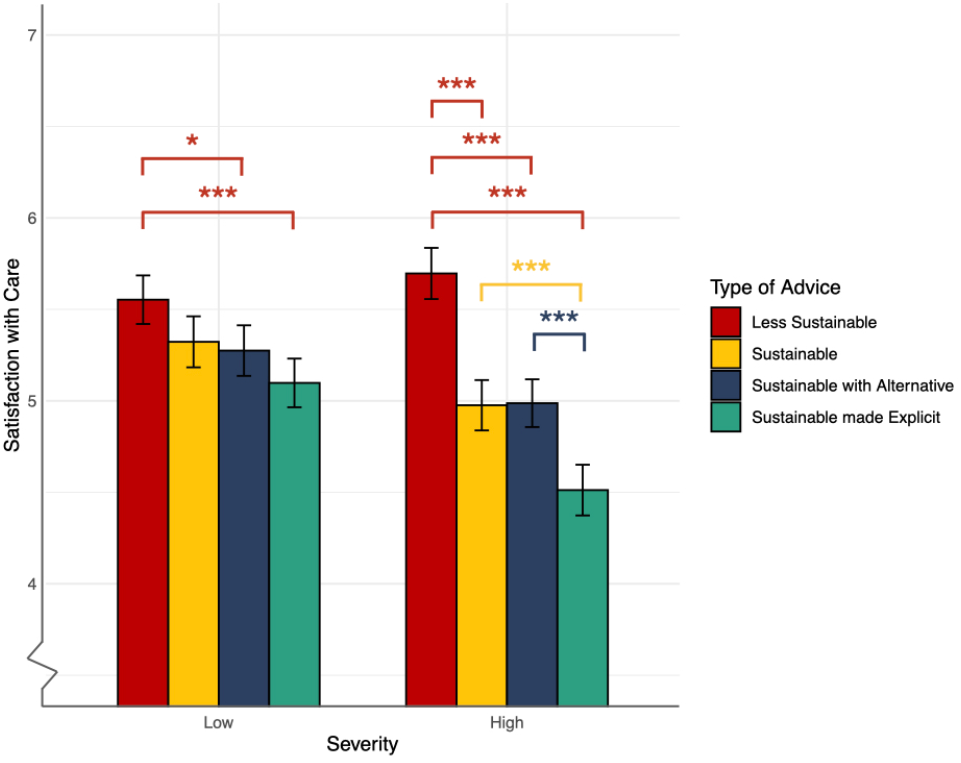
Participants’ satisfaction with care according to Severity and Type of Advice Note: * P<.05, ** P≤.01, *** P≤.001. Satisfaction with care was measured on seven-point Likert scales (1= strongly disagree, 7 = strongly agree).

The two-way interactions were further qualified by a three-way Type of Advice x Severity x Medical Problem interaction (P<.001), indicating that the aforementioned Type of Advice x Severity interaction pattern differed across medical problems. To decompose this effect, we tested the interaction for each of the medical problems separately. We found significant Type of Advice x Severity interactions for the pulmonary, locomotor, cardiovascular, and abdominal problems (P’s<.001). For the fatigue problem, the interaction was non-significant (P=.65).

Medical problems with significant interactions were decomposed further (Figure 3), again by comparing the effect of different types of advice for high- and low-severity conditions separately. Within the low-severity conditions, generally no effect of Type of Advice on satisfaction with care was found, except for the low-severity locomotor problem. For this vignette, the Less Sustainable advice yielded significantly higher scores compared to all other types of advice (P’s<.001). In the high-severity conditions, differences between the three Sustainable types of advice and the Less Sustainable advice were significant and large for the locomotor problem (P’s<.001) and significant but less pronounced for the cardiovascular and abdominal problems (P’s≤.002). Moreover, the pulmonary and abdominal problems yielded significantly lower satisfaction with care scores for the Sustainable made Explicit advice compared to the other Sustainable types of advice (P’s<.001). Further results are reported in Supplement E.

**Figure 3.**
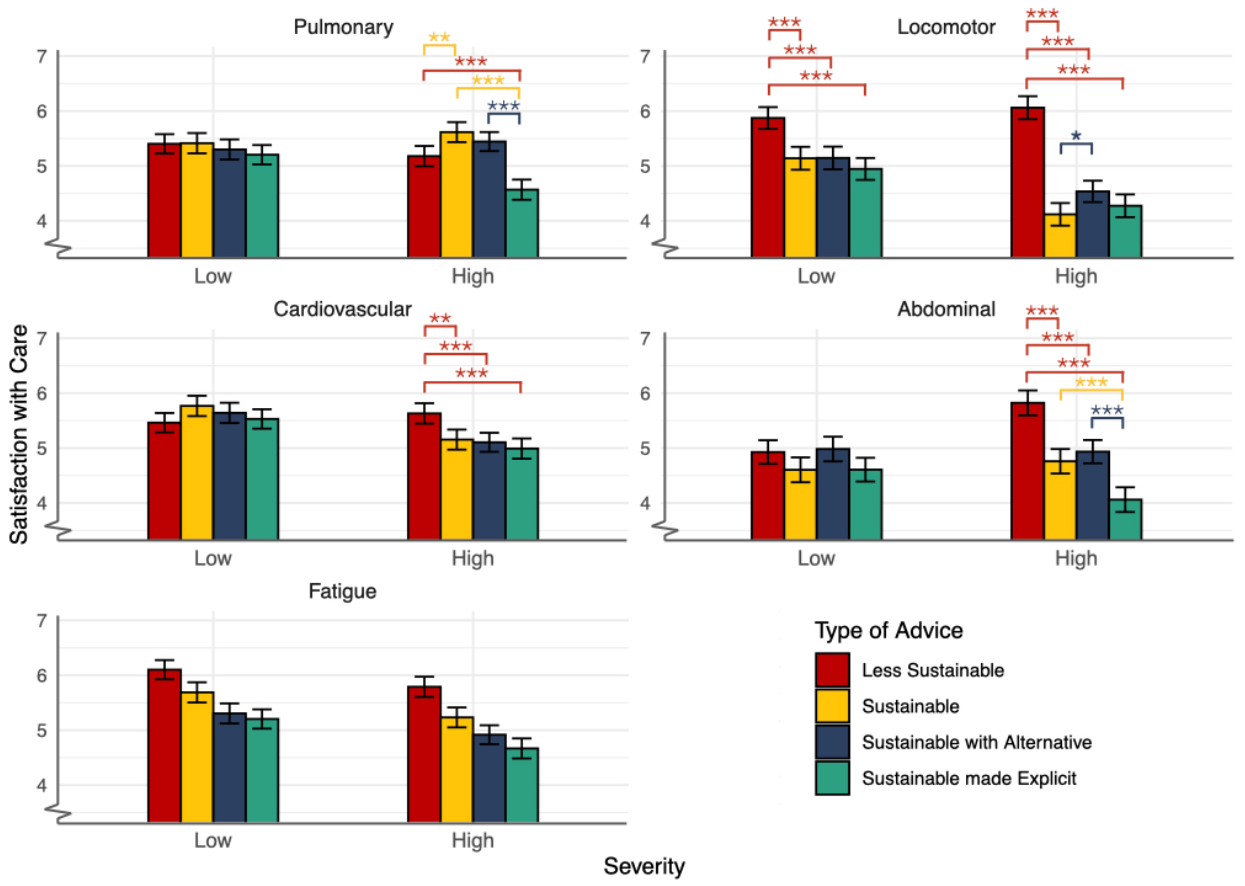
Participants’ satisfaction with care according to Severity and Type of Advice, split per medical problem Note: * P<.05, ** P≤.01, *** P≤.001. Pairwise comparisons for the low- and high-severity Fatigue vignette were not performed in the absence of a Type of Advice x Severity interaction. The trends for the fatigue problems within both the low- and high-severity groups follow the same trend as the general pattern displayed in Figure 2. Detailed results and 95%CIs are reported in Supplement E. Satisfaction with care was measured on seven-point Likert scales (1= strongly disagree, 7 = strongly agree).

## DISCUSSION

### Principal findings

In this randomised, double-blinded, experimental vignette study, we explored the effect of advising environmentally sustainable treatment options in clinical counselling with or without explicitly mentioning sustainability as an argument, compared to less sustainable (standard) treatment options – accounting for differences in severity and different types of medical problems. We found that participants’ satisfaction with care was significantly lower for the more sustainable treatment options than for the less sustainable options. This difference was primarily driven by the high-severity conditions and varied across medical problems. Moreover, we found that explicitly mentioning environmental sustainability in clinical counselling generally resulted in the lowest satisfaction with care scores; this difference was again mainly significant for some of the high-severity conditions.

### Contextualisation of findings

When specifically looking at the low-severity conditions, our findings largely corroborate Visser’s preceding primary care study.(12) That study found no difference in satisfaction with care between more sustainable and less sustainable treatment options. Notably, this included a similar vignette of prescribing asthma inhalers. In contrast to Visser’s study, we did find a significantly lower satisfaction with care for the locomotor problem – possibly reflecting participants’ expectation of physical follow-up as standard care or doubts regarding their self-efficacy to use eHealth, based on open question responses (Supplement C). Overall, these results indicate that a general practice setting, wherein problems may more frequently be perceived as low-severity, may be suitable for considering environmental sustainability in clinical counselling.

Results for our high-severity conditions indicated that advising sustainable treatment options leads to somewhat lower satisfaction with care in most cases. Recent hospital studies among gynaecology and cardiology patients indeed reported a lower willingness to consider the environmental impact of care for more severe medical problems.(13,16,17) However, in one of the gynaecology studies up to 62% of patients self-reported that they would generally choose the more sustainable treatment option in case of clinical equipoise.(13) This contrasting finding compared to our results may be explained by social desirability bias or overestimation of climate conscious decisions when patients self-report willingness in non-blinded designs.(20,21) Additionally, the contrast may be explained by patients’ underlying preference for the specific less sustainable standard treatment options in our study - due to familiarity, or the feeling that their health is not the (only) priority.

Further comparison with the literature is limited to general surveys inquiring participants’ willingness to consider the environmental impact of their treatment. Reportedly, 64% of the UK’s general population and 73% of a Dutch patient panel were willing to do so.(22,23) Once more, it seems contradicting that we observed a lower satisfaction with care for some high-severity medical problems when environmental sustainability was mentioned explicitly. Whilst multiple interpretations are possible (e.g. methodological differences, bias, the way environmental sustainability is addressed in the conversation) one may conclude that more medical problem-specific studies are necessary to understand whether and how environmental sustainability should be discussed and context-tailored in clinical counselling. Possibly, explicitly mentioning environmental sustainability has a stronger negative effect in high-severity conditions concerning a choice between two medical treatments (as observed in the pulmonary and abdominal problems in our study), as opposed to choices concerning clinical follow-up (the locomotor, cardiovascular, and fatigue problems).

### Research implications

Our study suggests that, primarily for low-severity conditions, advising more sustainable treatment options could be an acceptable way to integrate environmental sustainability in clinical counselling. However, it also suggests that explicitly discussing sustainability does not benefit patient satisfaction and can affect it negatively, mainly for high-severity conditions. Ethically, clinicians may therefore question whether it is ever warranted to explicitly discuss the environmental impact of a treatment. Contrasting opinions exist in the literature. Those in favour of ‘green informed consent’ stress the importance of a proactive approach to elicit patients’ preferences in respect of their potential ecological values and to fulfil clinicians’ social responsibility to promote environmentally friendly healthcare.(24-26) Those against explicitly discussing sustainability, argue that information on the environmental impact of treatments can be persuasive and potentially erodes the patient’s autonomous decision-making or the patient-provider relationship.(27,28)

Considering the generally less disputed notion that clinical care inherently needs to become more sustainable,(1,9,29) we argue that participants’ somewhat lower satisfaction scores for mainly the high-severity conditions in this study need not be a counterargument to environmentally sustainable counselling per se. Mean scores still signified a positive response to the underlying trust and acceptability statements. Moreover, while we considered sustainable and less sustainable treatments to be sufficiently equal in value, participants may not necessarily have shared the same perception based on the brief information provided.(30,31) Differences between treatments could be influenced by multiple factors (including participants’ expectation of standard care). Physicians’ needs may therefore be further clarification and nuance under which conditions (and in what form) environmentally sustainable counselling affects patients’ trust. Furthermore, if trust is affected, the question may be when such a decline becomes too large and what level of environmental benefit justifies it.

From a practical point of view, our findings suggest that clinicians’ situational awareness is key to understand when environmentally sustainable counselling may have undesired effects. A recent focus group study including patients and physicians suggested a “delicate balance between informing and burdening”.(14) Our results add that this balance may also differ according to the context, differ per health condition, and be most pronounced for severe health concerns.

### Strengths and limitations

Our study was a randomised, double-blinded vignette study measuring how participants reacted to environmentally sustainable treatment options and the integration of environmental impact in (hypothetical) clinical counselling. This design decreased the risks associated with self-reporting, such as social desirability bias and overestimation of climate-conscious behaviour. Furthermore, it was the first study of its kind to include a diverse set of health problems varying in severity, supporting the differentiation of findings to patients and healthcare professionals in different clinical settings.

Some limitations merit emphasis. First, we recruited a representative sample of the general Dutch population, rather than patients with the described medical problems. This choice may have limited participants’ ability to fully grasp the complexity and emotional aspects of the situations described. However, part of the participants indicated personal experience with the vignettes’ medical problems (in total, 71.4% had experienced at least one of the medical problems). Moreover, we regard ‘healthy’ participants to still offer valuable insights into patient perspectives in this novel research field. Second, we used experimental vignettes to measure differences between groups.

Vignette studies are considered a valid approach to identify drivers of variation in quality of care studies,(18,32) yet their approximation of more extensive, face-to-face counselling may yield different satisfaction with care outcomes than a real-life study. Notwithstanding, we aimed to study the relative differences between groups rather than absolute scores for satisfaction with care and formulated our conclusions accordingly. Future clinical studies may therefore better measure how high or low patients’ satisfaction with care is, when exposed to environmentally sustainable treatment options in real-life clinical counselling.

### Conclusions

Our findings suggest that advising more sustainable treatment options for more severe medical problems may negatively affect patients’ satisfaction with care, especially when sustainability is mentioned explicitly. However, the presence and size of the observed effect varied across medical problems and was generally non-significant or small for less severe medical problems. Future research should confirm our findings in more extensive real-life counselling and clarify how the context and form of environmentally sustainable counselling impact such an effect. Considering previous reports of patients’ interest in the environmental impact of healthcare, we currently call upon clinicians’ situational awareness to sense whether environmental sustainability should be discussed, or primarily guaranteed through system and institutional level decisions.

**What is already known on this topic**

- Integrating environmental sustainability in healthcare decision-making is suggested as a key strategy to achieve greener healthcare practices.
- Patients are generally interested in the environmental impact of healthcare and self-report to be willing to consider it in their treatment choices.
- Patients’ acceptance of involving environmental outcomes in shared decision-making varies and they may consider it an unconventional topic.

**What this study adds**

- Advising more sustainable treatment options for severe medical problems may generally have a negative effect on patients’ satisfaction with care, especially when sustainability is mentioned explicitly.
- The presence and potency of this effect varies per medical problem and should therefore be considered on a problem- and context-tailored basis.
- For less severe medical problems, advising more sustainable treatment options generally does not affect patients’ satisfaction with care.

## ETHICS STATEMENTS

### Ethical approval

Ethical approval was granted by the departmental review committee of the Leiden University Medical Center (#24-3033). No objection was made to the execution of the study by the institutional review board, as the Medical Research Involving Human Subjects Act (in Dutch: WMO) was confirmed not to apply to the study. Participants gave informed consent prior to participating in the vignette study and had the possibility to opt out after having received the debriefing upon completion of the survey.

### DATA AVAILABILITY STATEMENT

All materials related to this study, including anonymised patient data and the statistical code used in the analysis, are available from the Open Science Framework (<link> will be added upon publication </link>).

## FOOTNOTES

### Contributors

All authors collaboratively conceived the study, obtained funding, and developed the research design. EvB was in the lead for the planning, data collection, data analysis, interpretation of findings, and writing of the first and subsequent versions of the manuscript. LvG and MA closely supported the data analysis, interpretation of findings, and co-wrote the manuscript. EV, JA, and EB gave input for the interpretation of findings and supported the writing of the manuscript. All authors accept full responsibility for the work and conduct of the study, had access to the data, and controlled the decision to publish. The corresponding author attests that all listed authors meet authorship criteria and that no others meeting the criteria have been omitted.

### Transparency

The lead author (EvB) affirms that the manuscript is an honest, accurate, and transparent account of the study being reported; that no important aspects of the study have been omitted; and that any discrepancies from the study as preregistered have been explained.

### Dissemination to participants and related patient and public communities

We plan to share the findings of this study with (non-)governmental organisations involved in healthcare and will take considerable effort to share and discuss the relevance of our findings with the Netherlands Patient Federation and their subcommittees. We will also discuss our findings with clinicians by presenting at relevant events, publishing complementary blogs and/or non-specialist articles, and using social media opportunities.

