## Supplementary Files A-E for "Integrating environmental sustainability in clinical counselling: a randomised, double-blinded, experimental vignette study of satisfaction with care"

Table of contents:

| | | |
|---|---|---|
| **Supplement A** | Overview of experimental vignettes | pages 2–6 |
| **Supplement B** | Pilot study of experimental vignettes | pages 7–8 |
| **Supplement C** | Qualitative analysis of open-ended questions | pages 9–12 |
| **Supplement D** | Exploratory analysis of post-vignette statements | page 13 |
| **Supplement E** | Detailed results of statistical analysis and deviations from preregistration | page 14 |

### Supplement A: Overview of experimental vignettes

**Vignette design**
The medical problems and types of advice detailed below appeared in the vignettes, with the only difference that they were included in Dutch rather than in English. An indication of the low-severity and high-severity versions, the source of the scenario, and the different types of advice was included. Types of advice are as detailed in the manuscript: A=less sustainable, B=sustainable, C=sustainable with alternative, and D=sustainable made explicit.
The Dutch versions of the vignettes are available from the corresponding researcher upon request.

**1. Pulmonary**
Low-severity: asthma treatment
Based on: guideline 'Asthma in adults' of the Dutch Society of General Practitioners (*Nederlands Huisartsen Genootschap*).

*Scenario*:
You are visiting your general practitioner because you have a cough and have been feeling somewhat short of breath since a couple of weeks. After the conversation and an examination, the general practitioner concludes that you have asthma. The general practitioner discusses with you what to do next.

*Types of Advice*:
A) He/she proposes to prescribe you a metered dose inhaler, to make sure that you don't become short of breath again.
B) He/she proposes to prescribe you a dry powder inhaler, to make sure that you don't become short of breath again.
C) Different types of medication are available: 1) a metered dose inhaler, or 2) a dry powder inhaler.
He/she proposes to prescribe you the dry powder inhaler, to make sure that you don't become short of breath again.
D) Different types of medication are available: 1) a metered dose inhaler, or 2) a dry powder inhaler.
He/she proposes to prescribe you the dry powder inhaler, to make sure that you don't become short of breath again and because this medication is less burdensome for the environment than the metered dose inhaler.

High-severity: lung cancer treatment
Based on: guideline 'Non-small cell lung carcinoma' of the Dutch Society of Pulmonologists (*Nederlandse Vereniging van Artsen voor Longziekten en Tuberculose*) and LCA-research comparing video-assisted thoracic surgery and stereotactic body radiotherapy for early-stage NSCLC. [1]

*Scenario*:
Your general practitioner has referred you to the hospital because you have been coughing a lot and feeling tired for a couple of weeks. After the conversation and an examination, the physician concludes that you have early-stage lung cancer. The physician discusses with you what to do next.

*Types of Advice:*
A) He/she recommends surgery to remove the sick part of your lung.
B) He/she recommends targeted radiation to kill the cancer cells.
C) Two treatments are possible: 1) surgery to remove the sick part of your lung, or 2) targeted radiation therapy to treat the sick part of your lung. He/she recommends targeted radiation to kill the cancer cells.
D) Two treatments are possible: 1) surgery to remove the sick part of your lung, or 2) targeted radiation therapy to treat the sick part of your lung. He/she recommends targeted radiation to kill the cancer cells, also because radiotherapy is less burdensome for the environment than a surgery.

**2. Locomotor**
Low-severity: ankle distortion follow-up
Based on: current practice of the website *Thuisarts.nl* and the guideline "Ankle ligament injury" of the Dutch Society of General Practitioners (*Nederlands Huisartsen Genootschap*).

*Scenario:*

You are visiting your general practitioner because your ankle is hurting since a fall earlier today. After the conversation and an examination, the general practitioner concludes that you have a sprained ankle. The general practitioner discusses with you what to do next.

*Types of Advice:*
A) He/she proposes to make a follow-up appointment in one week and gives you movement exercises for the time in between, to make sure that your ankle recovers well.
B) He/she proposes not to make a follow-up appointment and to practice by yourself using the advice on the website "Thuisarts.nl", to make sure that your ankle recovers well.
C) Two different options are available: 1) a follow-up appointment in one week and movement exercises for the time in between, or 2) no follow-up appointment and practicing by yourself using the advice on the website "Thuisarts.nl". He/she proposes not to make a follow-up appointment and to practice by yourself, to make sure that your ankle recovers well.
D) Two different options are available: 1) a follow-up appointment in one week and movement exercises for the time in between, or 2) no follow-up appointment and practicing by yourself using the advice on the website "Thuisarts.nl". He/she proposes not to make a follow-up appointment and to practice by yourself, to make sure that your ankle recovers well and because it is better for the environment if you do not have to come to the GP's practice.

*NB: the website 'Thuisarts.nl' is a national, generally well-known website by the Dutch Society of General Practitioners which offers patients (and the general public) information about common complaints in general practice, advice for self-help or over the counter medication (e.g. paracetamol as a painkiller), and details when and for which complaints to seek medical advice from your own general practitioner.*

<u>High-severity: ankle fracture follow-up</u>
Based on: current use of the Virtual Fracture Care app for direct discharge in several Dutch hospitals for minor ankle fractures (Weber A or stable Weber B fractures) and the guideline "Ankle fractures" by the Dutch Society of Surgeons (*Nederlandse Vereniging voor Heelkunde*). [2]

*Scenario:*
Your general practitioner has referred you to the hospital because your ankle is hurting a lot since a fall earlier today and you cannot walk properly. After the conversation and an examination, the physician concludes that you have a broken ankle. The physician discusses with you what to do next.

*Types of Advice:*
A) He/she proposes to put a cast around your lower leg and wants to see you for a follow-up appointment at the hospital for further explanation and exercises, because this treatment will help the broken bone to heal properly.
B) He/she proposes to wear a brace and use an app on your phone which contains further explanation and exercises which you can do at home, without a follow-up appointment at the hospital, because this treatment will help the broken bone to heal properly.
C) Two treatments are possible: 1) to put a cast around your lower leg and make a follow-up appointment at the hospital, or 2) to wear a brace and use an app on your phone which contains further explanation and exercises which you can do at home, without a follow-up appointment at the hospital. He/she proposes to wear the brace and use the app on your phone, because this treatment will help the broken bone to heal properly.
D) Two treatments are possible: 1) to put a cast around your lower leg and make a follow-up appointment at the hospital, or 2) to wear a brace and use an app on your phone which contains further explanation and exercises which you can do at home, without a follow-up appointment at the hospital. He/she proposes to wear the brace and use the app on your phone, because this treatment will help the broken bone to heal properly and because it is better for the environment if you do not have to come to the hospital.

**3. Cardiovascular**
<u>Low-severity: hypertension follow-up</u>
Based on: follow-up of patients enrolled for cardiovascular risk management in general practices in Leiden, using non-invasive measurements at home. [3] Side note: considering the short travel distance between their home and the GP's practice for the average Dutch patient, it may be unlikely that the possible need to produce an additional blood pressure monitor will outweigh the environmental impact of the avoided travel distance.

*Scenario:*

You are visiting your general practitioner because you have hypertension since half a year. After the conversation and an examination, the general practitioner concludes that your measurements show a good result. The general practitioner discusses with you what to do next.

*Types of Advice:*
A) He/she proposes to make an annual follow-up appointment with the general practice assistant to adequately monitor your health.
B) He/she proposes to use a blood pressure monitor to perform your measurements at home, which the general practice assistant can view at a distance to adequately monitor your health.
C) Two different options are available: 1) an annual follow-up appointment with the general practice assistant, or 2) using a blood pressure monitor to perform your measurements at home, which the general practice assistant can view at a distance. He/she proposes to perform the measurements at home to adequately monitor your health.
D) Two different options are available: 1) an annual follow-up appointment with the general practice assistant, or 2) using a blood pressure monitor to perform your measurements at home, which the general practice assistant can view at a distance. He/she proposes to perform the measurements at home to adequately monitor your health and because it is better for the environment if you do not have to come to the GP's practice.

<u>High-severity: angina follow-up</u>
Based on: the standard protocol follow-up of patients post-myocardial infraction at the Leiden University Medical Centre by using non-invasive home measurements to replace some of the physical appointments at the hospital, avoiding the need for patients to travel to the hospital. [4] Side note: although the current scenario does not represent an actual myocardial infarction, we have chosen an approximately similar follow-up.

*Scenario:*
Your general practitioner has referred you to the hospital because you have been experiencing an occasional squeezing chest pain since a couple of weeks. After the conversation and an examination, the physician concludes that you have angina: a narrowing of a blood vessel to the heart, occasionally causing the heart not to get enough oxygen. The physician discusses with you what to do next.

*Types of Advice:*
A) He/she prescribes you medication and proposes to make a follow-up appointment including measurements at the hospital, so he/she can adequately measure your health.
B) He/she prescribes you medication and proposes that you measure your blood pressure at home and make recordings of your heart activity using a special watch. The physician can then view the results at a distance and schedule a video call, so he/she can adequately measure your health.
C) Two different options are available: 1) you get medication and a follow-up appointment including measurements at the hospital, or 2) you get medication, measure your blood pressure at home, and make recordings of your heart activity using a special watch. The physician can then view the results at a distance and schedule a video call. He/she proposes to perform the measurements at home, so he/she can adequately measure your health.
D) Two different options are available: 1) you get medication and a follow-up appointment including measurements at the hospital, or 2) you get medication, measure your blood pressure at home, and make recordings of your heart activity using a special watch. The physician can then view the results at a distance and schedule a video call. He/she proposes to perform the measurements at home, so he/she can adequately measure your health and because it is better for the environment if you do not have to come to the hospital.

**4. Abdominal**
<u>Low-severity: cholecystolithiasis treatment</u>
Based on: the guideline "Gallstones" of the Dutch Society of Surgeons (*Nederlandse Vereniging voor Heelkunde*) and research regarding the optimisation of indications for surgical intervention among patients with symptomatic (uncomplicated) cholecystolithiasis, including a decision aid. [5,6]

*Scenario:*
You are visiting your general practitioner because you have abdominal pain since today. After the conversation and an examination, the general practitioner concludes that you have gallstones. The general practitioner discusses with you what to do next.

*Types of Advice:*

A) He/she proposes to refer you to the hospital to have your gallbladder removed.
B) He/she proposes to wait, because the complaints will fade by themselves and because of the chance that the pain persists even after a surgery.
C) Two different options are available: 1) a referral to the hospital to have your gallbladder removed, or 2) wait for the complaints to fade. He/she proposes to wait, because the complaints will fade by themselves and because of the chance that the pain persists even after a surgery.
D) Two different options are available: 1) a referral to the hospital to have your gallbladder removed, or 2) wait for the complaints to fade. He/she proposes to wait, because the complaints will fade by themselves and because of the chance that the pain persists even after a surgery. Furthermore, waiting for the complaints to fade is less burdensome for the environment than a surgery.

<u>High-severity: appendicitis treatment</u>
Based on: the guideline "Acute appendicitis" of the Dutch Society of Surgeons (*Nederlandse Vereniging voor Heelkunde*), specifically referring to the treatment options of an uncomplicated appendicitis, [7] and ongoing care evaluation studies. Side note: to the best of our knowledge, there are no studies comparing the environmental impact of antibiotic (conservative) or surgical treatment of an uncomplicated appendicitis. In this case, we assumed that the environmental impact of conservative treatment would be lower, considering the incurred material and energy use of a surgery (including the possibility that for a certain percentage of patients a recurrence may occur).

*Scenario:*
Your general practitioner has referred you to the hospital because you have a strong abdominal pain since today. After the conversation and an examination, the physician concludes that you have an inflammation of your appendix. The physician discusses with you what to do next.

*Types of Advice:*
A) He/she proposes to remove the appendix via surgery, to make sure that the inflammation disappears.
B) He/she proposes to treat the inflammation with antibiotics in the hospital, to make sure that the inflammation disappears.
C) Two different options are available: 1) to remove the appendix via surgery, or 2) to treat the inflammation with antibiotics in the hospital. He/she proposes to treat you with antibiotics, to make sure that the inflammation disappears.
D) Two different options are available: 1) to remove the appendix via surgery, or 2) to treat the inflammation with antibiotics in the hospital. He/she proposes to treat you with antibiotics, to make sure that the inflammation disappears and because it is less burdensome for the environment than a surgery.

**5. Fatigue**
<u>Low-severity: possible hyperthyroidism work-up</u>
Based on: the guideline "Thyroid disease" of the Dutch Society of General Practitioners (*Nederlands Huisartsen Genootschap*). Side note: considering the short travel distance between their home and the GP's practice for the average Dutch patient, it may be unlikely that a substantial environmental benefit is obtained in terms of avoided travel distance.

*Scenario:*
You are visiting your general practitioner because in the last two weeks you are frequently tired and have lost some weight. After the conversation and an examination, the general practitioner says that he/she wants to perform additional blood tests. The general practitioner discusses with you what to do next.

*Types of Advice:*
A) He/she proposes that you make a follow-up appointment in a couple of days, so you can hear about the test results.
B) He/she proposes that you receive an online message (e-consultation) from the general practitioner in a couple of days, so you can hear about the test results.
C) Two different options are available: 1) a follow-up appointment in a couple of days, or 2) an online message (e-consultation) from the general practitioner in a couple of days. He/she proposes to send you an e-consultation, so you can hear about the test results.
D) Two different options are available: 1) a follow-up appointment in a couple of days, or 2) an online message (e-consultation) from the general practitioner in a couple of days. He/she proposes to send you an e-

consultation, so you can hear about the test results. Furthermore, it is better for the environment if you do not have to come to the GP's practice.

High-severity: possible malignancy work-up
Based on: the guideline "Primary tumor unknown" of the Dutch Society of Pathologists (*Nederlandse Vereniging voor Pathologie*). Side note: a "scan of the body" may be perceived as a rather non-specific or untargeted approach to primary tumor diagnostics, which in medical practice is frequently guided by anamnesis, medical history, and/or blood tests. Considering the aim to keep the text rather brief, the choice was made to omit any further information.

*Scenario:*
Your general practitioner has referred you to the hospital because in the last two weeks you are very tired and have lost a lot of weight. After the conversation and an examination, the physician says that he/she wants to make a scan of the body for further diagnostics. The physician discusses with you what to do next.

*Types of Advice:*
A) He/she proposes that you and a family member come for a follow-up appointment in one week, so you can discuss the results together.
B) He/she proposes that you and a family member make an appointment via video calling in one week, so you can discuss the results together.
C) Two different options are available: 1) a follow-up appointment in one week, with a family member, or 2) a follow-up appointment via video calling in one week, which a family member can join. He/she proposes to video call, so you can discuss the results together.
D) Two different options are available: 1) a follow-up appointment in one week, with a family member, or 2) a follow-up appointment via video calling in one week, which a family member can join. He/she proposes to video call, so you can discuss the results together and because it is better for the environment if you do not have to come to the hospital.

### Supplement B: Pilot study of experimental vignettes

#### Pilot study design

Prior to the main study, we conducted a pilot study where we tested four sets (i.e. textual variations) of five vignettes for their perceived severity and urgency,* and asked respondents to comment on their readability. The study had a 2 Severity (low vs high) x 2 Urgency (low vs high) between-subjects design and participants were randomly allocated to just one set of vignettes (e.g. low-severity and low-urgency). For this pilot study, we aimed to recruit 80 participants, which we deemed sufficient to compare the different versions of the vignettes.

* Considering the minimal textual differences between the low- and high-urgency versions of vignettes and the choice not to use them in the final study (due to the absence of a clear differentiating effect), the vignettes are not included in this supplementary file. To illustrate the difference, low-urgency pulmonary scenarios (both low- and high-severity) would e.g. read:

*"You are visiting your general practitioner because you have a cough and have been feeling somewhat short of breath since a couple of weeks. After the conversation and an examination, the general practitioner concludes that you have asthma. The general practitioner discusses with you what to do next."*

*"Your general practitioner has referred you to the hospital because you have been coughing a lot and feeling tired for a couple of weeks. After the conversation and an examination, the physician concludes that you have early-stage lung cancer. The physician discusses with you what to do next."*

Alternatively, the high-urgency scenarios would read:

*"You are urgently visiting your general practitioner because you have had a cough for a couple of months and have started to feel somewhat short of breath recently. After the conversation and an examination, the general practitioner concludes that you have asthma. The general practitioner discusses with you what to do next."*

*"Your general practitioner has directly referred you to the hospital because you have been coughing a lot for a couple of months, recently including traces of blood. After the conversation and an examination, the physician concludes that you have early-stage lung cancer. The physician discusses with you what to do next."*

We recruited a sample of Dutch-speaking participants via Prolific Academic. Participants were presented with the respective vignettes in random order and asked to rank the perceived severity and urgency of the described situation on two seven-point Likert scales (1=not severe/urgent, 7=very severe/urgent). In addition, participants provided their age, gender, education level, and self-rated health status (operationalised as a single SF-36 item, similar to the main study) and answered an open question about the readability of the vignettes. Participants gave informed consent prior to answering the vignettes. Similar to the main study, we only informed patients that we wanted to get their opinion regarding hypothetical doctor's visits. Once they had completed all the questions, they received a short debriefing.
To compare the scores on perceived severity and urgency for the different versions of the vignettes descriptive statistics were used (i.e. no statistical testing of differences was performed).

#### Pilot study results

In total, 85 participants completed the pilot study. Compared to the sample of the main study, participants were relatively young, highly educated, and generally had a "(very) good" self-rated health status (Table B1).

*Table B1. Sample descriptives based on allocated group*

| | **Severity, low** | | **Severity, high** | | **Pooled** |
|---|---|---|---|---|---|
| | **Urgency, low** | **Urgency, high** | **Urgency, low** | **Urgency, high** | |
| | N = 21 | N = 22 | N = 21 | N = 21 | N = 85 |
| | N (%) | N (%) | N (%) | N (%) | N (%) |
| Age, mean [SD] | 34 [14] | 30 [8] | 36 [10] | 29 [8] | 32 [10] |
| Gender | | | | | |
| Male | 10 (48) | 9 (41) | 14 (67) | 9 (43) | 42 (49) |
| Female | 11 (52) | 12 (55) | 7 (33) | 12 (57) | 42 (49) |
| Other | 0 (0) | 1 (5) | 0 (0) | 0 (0) | 1 (1) |
| Education level | | | | | |
| Low | 1 (5) | 2 (9) | 3 (14) | 3 (15) | 9 (11) |
| Middle | 6 (29) | 5 (23) | 5 (24) | 5 (24) | 21 (25) |
| High | 14 (67) | 15 (68) | 13 (62) | 13 (62) | 55 (65) |
| Self-rated health status | | | | | |
| Poor | 0 (0) | 0 (0) | 0 (0) | 0 (0) | 0 (0) |
| Fair | 3 (14) | 3 (14) | 6 (29) | 5 (24) | 17 (20) |
| Good | 9 (43) | 13 (59) | 9 (43) | 7 (33) | 38 (45) |
| Very good | 7 (33) | 3 (14) | 6 (29) | 6 (29) | 22 (26) |
| Excellent | 2 (10) | 3 (14) | 0 (0) | 3 (14) | 8 (9) |

For perceived severity (Table B2), we generally observed large and consistent mean differences between the low- and high-severity conditions. The smallest mean difference was observed for the abdominal problem, possibly due to a ceiling effect (i.e., the severity scores were already relatively high in the low-severity condition). Still, this mean difference was approximately 0.6 and in the same direction as the other differences. For perceived urgency (Table B3), observed differences were often negligible and not robust (i.e. observed differences were also often in the opposite direction).

*Table B2. Perceived severity scores per allocated group:*

| **Medical Problem** | **Severity, low** | | **Severity, high** | |
|---|---|---|---|---|
| | **Urgency, low (N = 21)** Mean [SD] | **Urgency, high (N = 22)** Mean [SD] | **Urgency, low (N = 21)** Mean [SD] | **Urgency, high (N = 21)** Mean [SD] |
| **Pulmonary** | 4,14 [1.49] | 3.95 [1.21] | 6.76 [0.44] | 6.52 [0.75] |
| **Locomotor** | 2.48 [1.12] | 3.00 [1.20] | 4.29 [1.35] | 4.52 [1.44] |
| **Cardiovascular** | 3.00 [1.34] | 3.86 [1.58] | 5.48 [0.98] | 6.05 [0.97] |
| **Abdominal** | 4.90 [1.14] | 4.86 [1.25] | 5.48 [1.33] | 5.24 [1.26] |
| **Fatigue** | 3.90 [1.34] | 4.77 [0.92] | 4.86 [1.24] | 4.48 [1.57] |

*Table B3. Perceived urgency scores per allocated group:*

| **Medical Problem** | **Severity, low** | | **Severity, high** | |
|---|---|---|---|---|
| | **Urgency, low (N =21)** Mean [SD] | **Urgency, high (N = 22)** Mean [SD] | **Urgency, low (N = 21)** Mean [SD] | **Urgency, high (N = 21)** Mean [SD] |
| **Pulmonary** | 3.33 [1.35] | 3.68 [1.32] | 6.62 [0.59] | 6.48 [0.98] |
| **Locomotor** | 3.00 [1.49] | 3.00 [1.38] | 5.14 [1.42] | 4.95 [1.53] |
| **Cardiovascular** | 2.90 [1.67] | 3.64 [1.65] | 5.52 [1.08] | 6.00 [0.89] |
| **Abdominal** | 5.00 [1.34] | 5.18 [1.37] | 6.14 [1.11] | 6.05 [1.16] |
| **Fatigue** | 4.14 [1.85] | 4.50 [1.10] | 4.57 [1.33] | 4.52 [1.50] |

Consequently, we decided to select Severity as factor in the main study, continuing the differentiation between low- and high-severity medical conditions. We dropped the high-urgency textual variations of the vignettes.

The open question regarding readability yielded 34 responses (=40% response rate), indicating no problems with vignette and/or question readability.

### Supplement C: Qualitative analysis of open-ended questions

**Qualitative analysis – principal findings**
EvB inductively analysed the vignette-related open-ended questions using content analysis and qualitatively explored differences in themes between groups. Findings were discussed with the other co-authors during the interpretation of the Type of Advice x Severity interaction for the different medical problems and considered in the writing up of the manuscript.

In total, 1,104 participants (72%) responded to the physician's advice via the open-ended questions after the vignettes. Irrespective of group allocation, most participants asked the physician to elaborate on the advised treatment and its expected benefits/risks, recovery and prognosis, follow-up plans, and possibilities for lifestyle interventions or self-help to reduce or prevent further complaints. In the Less Sustainable condition, some questioned the need for additional diagnostics or inquired whether an alternative treatment was available - especially for surgical interventions.

In the Sustainable conditions, participants also expressed doubts regarding the proposed treatment, yet particularly when they differed from their expectation of standard care. This was mostly the case for the locomotor and abdominal conditions – i.e. eHealth-based follow up of injuries/fractures, like the Virtual Fracture Clinic, [1] and nonoperative treatment using antibiotics for uncomplicated appendicitis. [2] Moreover, some participants voiced concerns regarding their ability to use telemedicine, indicated that they would need more guidance to perform exercises or measurements correctly, or stated that they would prefer in-person visits.

In the Sustainable made Explicit group, approximately a quarter of participants addressed the inclusion of environmental impact in the physician's advice. Most of these participants stated that the environmental impact of a treatment should not be as important as their personal health or asked if the suggested treatment would be different when only considering health outcomes. A minority explicitly raised concern or dislike regarding the physician for including the environmental impact in their advice.

*NB: a detailed overview of the identified content categories is provided on the next pages. Categories have been listed by frequency per Type of Advice x Severity participant group (i.e. according to group allocation as visualised in Figure 1 in the manuscript). Those categories listed at the top of every list were coined by more participants than those listed at the bottom. Note that this does not indicate importance of the statements. In case a category specifically related to one of the medical problems, this was indicated.*

[1] Khan SA, Asokan A, Handford C, Logan P, Moores T. How useful are virtual fracture clinics?: a systematic review. Bone Jt Open. 2020;1(11):683-90.
[2] de Almeida Leite RM, Seo DJ, Gomez-Eslava B, Hossain S, Lesegretain A, et al. Nonoperative vs Operative Management of Uncomplicated Acute Appendicitis: A Systematic Review and Meta-analysis. JAMA Surg. 2022;157(9):828–834.

**Participants in the Less Sustainable condition**
High-severity:
- Questions regarding prognosis for recovery
- Follow-up questions related to medical advice (i.e. participant would like further information)
- Wonders what disease/condition the physician expects (*fatigue vignette*)
- Wonders if the suggested medication cures the disease/complaints
- Asks what the other treatment options are, wants more extensive counselling
- Wonders if the physician should do further diagnostic tests (*cardiovascular and abdominal vignette*)
- Wonders if they don't need surgery (*locomotor vignette*)
- Wonders what the consequences are of not taking the suggested treatment
- Wonders what follow-up will look like
- Wonders if they don't need an X-ray first
- Asks what they can do themselves (e.g. lifestyle)
- Wonders what the cause of the disease is (*pulmonary vignette*)
- In case of 'no abnormalities', can't further check-ups happen via phone?

Low-severity:
- Would prefer a referral to a medical specialist for further treatment (*pulmonary* + *abdominal vignette*)
- Questions whether it is necessary to remove the gall bladder. (*abdominal vignette*)
- Would like more extensive treatment counselling (e.g. do I really need surgery; what follow-up would the physician recommend; what side-effects to expect)
- Further diagnostics/second opinion required (*pulmonary* + *abdominal vignette*)
- Can we do check-ups every half year instead of every year? (*cardiovascular vignette*)
- Wants to know about the origin of the disease (*pulmonary vignette*)
- Wonders what they can do themselves to relieve complaints
- Question if additional treatment (e.g. physiotherapy) would be helpful
- Questions/suggestions what follow-up will look like
- Medication-related questions
- Don't I need an X-ray of the ankle? (*locomotor vignette*)
- Is follow-up for a sprained ankle really necessary?
- Further questions about the (danger of the) disease

**Participants in the Sustainable condition**
High-severity:
- Doubts whether the proposed treatment (e.g. no surgery) will be a good cure for the disease
- Follow-up questions related to the medical advice (i.e. what are the risks & prognosis)
- Prefers a physical appointment over a virtual one
- Wonders if they should have a check-up of the leg after follow-up completion (*locomotor vignette*)
- Follow-up questions related to medical advice (i.e. participant would like further information)
- Patient has a different expectation (i.e. I want an X-ray, I want a cast; *locomotor vignette*)
- Medication side-effects
- Wants to know why a virtual appointment is chosen
- What other options are available?
- Wonders if enough tests have been performed to arrive at the diagnosis (*pulmonary vignette*)
- Wants to know when to act themselves, rather than waiting for physician
- Question when the treatment will be evaluated and what the alternative will be
- For how long do I need telemonitoring? And shouldn't I have additional tests every now and then?

Low-severity:
- Patient has a different treatment suggestion (e.g. shouldn't I go to the physiotherapist)
- For how long will I need the treatment?
- Patient wonders if there is a different treatment option available
- Wants to know who will verify if the exercises are performed well (*locomotor vignette*)
- Asks for painkillers or pain relief advice
- Wants to know the origin of the disease
- Wonders if further diagnostics are required (e.g. X-ray)
- Patient expresses that the physician should tell them about an alternative (which they know exists)
- Wants to know about potential side-effects of the treatment
- Want to know by when complaints are expected to resolve (*abdominal vignette*)
- Wants to know how complaints can be gone all of a sudden (*cardiovascular vignette*)

- Digital results are fine, but expects a personal phone call if bad results or expects a clear layperson explanation in the online system (*fatigue vignette*)
- Patient wants to know when to measure and how to measure blood pressure (*cardiovascular vignette*)
- Does the telemonitoring device incur out-of-pocket costs?
- Wants to know what they could do themselves to reduce complaints
- Wants to know why telemonitoring is chosen over the other option

**Participants in the Sustainable with Alternative condition**
High-severity:
- Patient wonders how effective the treatment is and what the risks are (*abdominal vignette*)
- Question about success rate of the treatments (*pulmonary vignette*)
- Wants to know what happens with/based on the telemonitoring measurements
- Patient has a different treatment suggestion (surgery, referral, second opinion)
- Questions about risks of the advised treatment (e.g. telemonitoring,
- Wants more explanation why the chosen options is the preferred option
- Follow-up questions related to medical advice (i.e. participant would like further information)
- Questions about prognosis
- Preference for physical over virtual check-up
- Wonders if they can still get the other treatment if this one doesn't work (*abdominal vignette*)
- Wonders what to do in the meantime if complaints get worse
- Would like an X-ray in the follow-up (*locomotor vignette*)
- Would prefer a cast over a brace (*locomotor vignette*)

Low-severity:
- Wants more information why this treatment instead of the other is chosen
- Wants to know when they can contact the physician again if complaints haven't resolved
- Wants to know when/how the treatment will be evaluated
- Would like more information about recovery period and course of the suggested treatment
- Would like to discuss prognosis and what will happen if lab results are bad (*fatigue vignette*)
- Prefers a physical consultation (or explanation why virtual)
- Wonders what happens if telemonitoring results are bad ("what happens then")
- Would like additional pain killers
- Medication-related questions
- Wonders how to know if they do the exercises right (*locomotor vignette*)
- Don't I need an X-ray of the ankle?
- Will the telemonitoring device incur out-of-pocket costs?
- Would prefer surgery (*abdominal vignette*)
- Wants a referral to a medical specialist for further diagnostics (*pulmonary vignette*)
- Wish to avoid medication as much as possible (*cardiovascular vignette*)
- Indicates not to visit the GP for these types of complaints

**Participants in the Sustainable made Explicit condition**
High Severity:
- Can I still have the other treatment if the radiation doesn't work (*pulmonary vignette*)
- Questions about the reliability of telemonitoring (*locomotor vignette*)
- Wants more information about the reason that one treatment is preferred
- Questions about success rate and/or risks of suggested treatment (mainly *abdominal vignette*)
- Wonders what the best treatment is without considering the environment
- Concerns and/or expressed negative sentiment regarding the environmental information
- Prefers the alternative option/advice based on own experience or hear-say
- What are other patients' experiences with a broken ankle and brace? (*locomotor vignette*)
- Prefers a physical consultation
- "What is the *real* reason not to do a personal appointment with the results?" (*fatigue vignette*)
- Questions about prognosis

Low Severity:
- Prefers a physical consultation over a virtual one
- Questions regarding follow-up: when to contact, what will it look like, when/how the effect will be evaluated
- Questions what they could do themselves for treatment or prevention

- Further questions regarding differences between the two inhalers (*pulmonary vignette*)
- Requests for further counselling to decide which treatment to take (mainly *abdominal vignette*)
- Don't I need physiotherapy? (*locomotor vignette*)
- Low credibility or perceived irrelevance of environmental argument
- Wonders what the best treatment is without considering the environment
- Question whether the other treatment option would also be possible or is still available if the current advice does not work
- Suspects that there is an underlying reason that is not mentioned (time)
- Question what the physician would advise/which treatment they would choose themselves

### Supplement D: Exploratory analysis of post-vignette statements

#### Elaboration of methods

After the vignettes, all participants (irrespective of group allocation) were presented a short piece of information about environmental sustainability in healthcare and then asked to indicate their agreement (5-point Likert scale) with three related statements:

> Healthcare has an influence on nature and the environment. For example, due to energy use, production of waste, and commute by car. To reduce this influence, an increasing number of healthcare professionals are trying to practice 'sustainable healthcare'. Below are three questions related to healthcare and its influence on nature and the environment.
>
> *To what extent do you agree with the following statements?*
> (1= fully disagree; 5 = fully agree)
> 1. Considering nature and the environment suits the job of a doctor.
> 2. I expect it to negatively affect my health if my physician also considers nature and the environment.
> 3. I would prefer a treatment which I know to have a smaller influence on nature and the environment.

Next, participants were presented the question whether they believe that climate change exists (described in the manuscript) and when they responded positively, another three statements regarding their outlook on climate change:

> *To what extent do you agree with the following statements?*
> (1= fully disagree; 5 = fully agree)
> 1. I believe that climate change is a result of the way we, humans, live on this planet.
> 2. I worry about the consequences of climate change.
> 3. I believe that the world will be in a better place 10 years from now.

As appears from the statements above, we consistently phrased two statements in a positive way and one statement in a negative way (or vice versa). In case a composite score was calculated, we first reverse-coded participant scores.

#### Sustainable healthcare and climate change - results

For the three statements related to sustainable healthcare (Cronbach's α=.71), participants' composite scores varied between 3.3 [SD 0.9] and 2.7 [0.9] out of 5 (Table D1). Both groups that received the physician's advice including environmental sustainability (SmE groups) scored lowest. The Kruskal-Wallis test indicated significant between group differences (P<.01).

For the questions related to climate change, 5.5 to 12.4% (N=10–26) of participants (across groups) indicated not to believe in climate change. Those that did believe in climate change, generally indicated that: climate change is a consequence of human behaviour (4.0 [SD 1.0] to 4.3 [SD 0.8] out of 5), indicated some worry about the consequences of climate change (3.8 [1.1] to 4.0 [0.9]), and replied negatively to the statement that the world will look better in ten years (2.2 [0.9] to 2.4 [0.8]). The Kruskal-Wallis test only indicated a significant between group difference for the question regarding human behaviour (P=.03).

**Table D1**. Participants' scores for the sustainable healthcare and climate change statements (1-5 Likert scales)

| | **High severity** | | | | **Low severity** | | | |
|---|---|---|---|---|---|---|---|---|
| | **LS** | **S** | **SwA** | **SmE** | **LS** | **S** | **SwA** | **SmE** |
| | Mean [SD] | Mean [SD] | Mean [SD] | Mean [SD] | Mean [SD] | Mean [SD] | Mean [SD] | Mean [SD] |
| SH composite | 3.1 [0.9] | 3.1 [0.9] | 3.1 [0.8] | 2.7 [0.9] | 3.0 [0.8] | 3.3 [0.9] | 3.2 [0.9] | 2.8 [0.9] |
| SH Q1 (+) | 3.1 [1.1] | 3.1 [1.1] | 3.2 [1.0] | 2.8 [1.1] | 3.1 [1.0] | 3.3 [1.1] | 3.2 [1.1] | 2.8 [1.1] |
| SH Q2 (-) | 2.7 [1.2] | 2.8 [1.1] | 2.7 [1.1] | 3.2 [1.1] | 2.7 [1.0] | 2.5 [1.1] | 2.6 [1.1] | 3.0 [1.1] |
| SH Q3 (+) | 2.9 [1.1] | 2.8 [1.1] | 2.9 [1.0] | 2.6 [1.1] | 2.7 [1.0] | 3.0 [1.1] | 2.9 [1.1] | 2.7 [1.0] |
| CC Q1 (-) | 4.2 [0.9] | 4.1 [0.9] | 4.2 [0.9] | 4.3 [0.8] | 4.1 [0.9] | 4.1 [0.9] | 4.3 [0.8] | 4.0 [1.0] |
| CC Q2 (-) | 3.8 [1.0] | 4.0 [0.9] | 3.8 [1.1] | 3.9 [1.0] | 3.9 [0.9] | 3.8 [1.0] | 3.9 [1.0] | 3.8 [1.1] |
| CC Q3 (+) | 2.3 [0.9] | 2.2 [0.9] | 2.3 [1.0] | 2.4 [0.8] | 2.4 [0.8] | 2.4 [0.8] | 2.3 [0.8] | 2.3 [0.9] |

*Legend: LS = Less Sustainable, S = Sustainable, SwA = Sustainable with Alternative, SmE = Sustainable made Explicit, SH = sustainable healthcare, CC = climate change.*

### Supplement E: Detailed results of statistical analysis and deviations from preregistration

A detailed and transparent overview of the data preparation, exploratory analysis, and statistical analysis of the study aims is available as a .html R Markdown file via the Open Science Framework: <link added upon publication, directly available as .html file to reviewers during submission process>. In the corresponding sections, this file also contains an overview of any of the deviations from the preregistration on AsPredicted (aspredicted.org/mhzn-pmv6.pdf).

For transparency, the deviations are also listed separately directly below:

- **Regarding reliability testing**
  After re-discussing with the research team, we concluded that the three statements related to Climate Change measured different constructs instead of the same construct. E.g.: the first statement inquired participants' perspective whether climate change is caused by human activity, while the third statement inquired whether they had a positive outlook on the future. Therefore, we did not perform reliability testing on these statements.
- **Regarding randomisation check**
  Whereas this had not been made explicit in the preregistration, we regarded the questions answered after completion of the vignettes to contain one group of questions that were very unlikely to have been influenced by participants' exposure to the vignettes (i.e. the questions regarding their previous experience with the Medical Problems in the vignettes and whether they believed in climate change or not); and one group that could have been influenced by their exposure to the vignettes (i.e. statements regarding their perspective on Sustainable Healthcare and Climate Change). Therefore, we performed a randomisation check for the first group (considered sample characteristics), but not for the second group (considered as descriptives for exploratory analysis).
